# COGNITIVE ADVERSE EFFECTS OF LACOSAMIDE IN PATIENTS WITH LOCALIZATION RELATED EPILEPSY - A PROSPECTIVE OBSERVATIONAL STUDY

**DOI:** 10.1101/2021.05.13.21257196

**Authors:** Sophia B Modi, S Asha, Thomas Iype, G K Libu, Reeja Rajan

## Abstract

Newer antiepileptic drugs (AEDs) offer favourable safety profiles than the previously used AEDs. Despite the introduction of many AEDs, a large number of patients continue to suffer from uncontrolled partial-onset seizures which have considerable impact on a patient’s quality of life. Lacosamide (LCM) is a third generation AED approved for adjunctive use in partial-onset seizures. Patients with epilepsy frequently experience cognitive dysfunctions due to a variety of factors. Because AEDs are the major therapeutic modality for epilepsy, the adverse effects of AEDs on cognition are important.

**Objectives:** To assess the adverse effects of lacosamide on cognition among patients with localization related epilepsy to whom lacosamide is given as adjuvant therapy.

**METHODOLOGY:** An open labelled prospective observational study in 22 patients who suffered from localization related epilepsy.

**Results:** Average Initial seizure frequency per month was 3.56 (SD 2.58) and median frequency 2.5 seizures per month. Range being 1-8 per month. At the final followup at 6months, only 2 persons experienced seizure and that too only single episodes. The difference in frequency is statistically significant (Wilcoxon Signed Ranks TestP <0.001). All the pre and post lacosamide cognition scores showed statistically significant positive correlation in this study.

**Conclusion:** Excellent seizure control is observed in patients with refractory localization related epilepsy treated with lacosamide. Also, lacosamide has no serious adverse effects or drug interactions. In this study, it is observed that unlike many AEDs, lacosamide contributed to significant improvement in cognition and can improve the quality of life in such patients.

## Introduction

Epilepsy is one of the most common disorders of the brain affecting around 50 million people worldwide.[1] Lacosamide (LCM), a third generation antiepileptic drug approved for adjunctive use in partial-onset seizures has two novel mechanisms of action and favourable pharmacokinetic and safety profiles. [2,3,4,5]

More than 30% of epilepsy patients remain refractory to pharmacotherapy despite the advent of new AEDs over last two decades. [6] Because AEDs are the major therapeutic modality for epilepsy, the adverse effects of AEDs on cognition are important. [7] Previous studies on lacosamide [2,8,9,10,11] observed that cognitive adverse effects (memory impairment, confusional state and disturbance in attention) were minimal and dose dependent.

The aim of this study was to obtain reliable data on cognitive effects of lacosamide in Indian patients with localization related epilepsy.

### Objectives

- To assess the adverse effects of lacosamide on cognition among patients with localization related epilepsy to whom lacosamide is given as adjuvant therapy.
- To study the seizure control profile of lacosamide in localization related epilepsy.

## METHODOLOGY

This open labelled prospective observational study was done at Government Medical college, Thiruvananthapuram. Patients were recruited from epilepsy patients attending the Neurology OPD from January 2012 to December 2012. We recruited patients with drug refractory localisation related epilepsy who were given an adequate trial of at least two AEDs to maximum tolerated dose. Patients from both genders were included who had seizures of more than 6 months duration with a seizure frequency of at least 2 seizures in previous 3 months. We excluded patients who were pregnant, had renal or hepatic dysfunction, patients with pre-existent cognitive dysfunction, with psychosis, alcohol& substance abuse. We also excluded patients with active suicidal plan/intent or active suicidal thoughts in the last 6 months and those with prior history of cardiac arrhythmia.

### Sample size and selection

According to the prescription rate of lacosamide at the study setting, we expected a sample size of 30. Only 25 patients were prescribed lacosamide during the study period. Of them, 22 satisfied the inclusion criteria and they were studied.

### STUDY TOOLS

- Pre tested Proforma
- Malayalam adaptation of Addenbrooke’s cognitive examination (M-ACE)
- Digit backward memory span
- Block design
- Digit symbol Test
- Trail making test-part A
- Engel’s Seizure Scoring System
- Naranjo’s score
- Hospital Anxiety Depression Score

Ethics committee clearance was obtained, informed consent was obtained from each subject / guardian/ relative and confidentiality and anonymity of the patient’s information were maintained

### Data collection

This study comprehended minimum 8 visits- an initial visit, follow up visits every 2 weeks in the initial 2months and monthly follow up visits for the next 4 months. Every subject was followed up at least for a period of 6 months after reaching the maintenance dose of lacosamide.

At the first visit, socio demographic and clinical details were collected. Baseline evaluation of the seizure type and frequency was done. Baseline cognition was assessed. Engel system was used to score seizure burden at the first visit. Following these baseline assessments, lacosamide was prescribed.

At the second to seventh OPD visit, assessment of vital signs and weight followed by physical and neurologic examinations, seizure frequency, and adverse effects were done. Engels scoring and Naranjo’s scoring were also done.

On the eighth OPD visit, apart from all other routine tests, battery of cognitive tests was repeated.

Collected data were entered in excel and analyzed using SPSS16. Relevant variables were expressed as means and standard deviation, medians, proportions and their 95%CI, appropriately.

Appropriate statistical tests (Wilcoxon Signed Ranks Test, Mann-Whitney U, Kruskal-Wallis Test, One sample t test, Chi-square test, Fisher’s exact, Spearman’s correlation etc) were performed to find out the statistical significance of changes in the cognitive variables and association with other variables studied.

## RESULTS AND DISCUSSION

Average age of the study population was 36.05. Majority were males (63.6%). 59.1% of the subjects were socially active. 31.8% had high school education and 27.3% had higher education. Most of the study subjects belonged to BPL category (81.8%). Family history of seizure disorder was reported by 2 (9%) subjects.

### Baseline characteristics

In the present study, the most common time of onset of epilepsy was adolescence (31.8%) the next being childhood (27.3%) and mean duration of epilepsy in the subjects was 20.8 years. Average baseline seizure frequency per six months was 21.4. Aura was present only in 23% of patients. 27.3% subjects had EEG abnormalities and 45.5% had CT/MRI abnormalities.

Most of the patients in this study were receiving 3 AEDs (59.1%) concomitantly. In this study, 16 (72.7%) subjects continued lacosamide till final follow up. Lacosamide was discontinued by 27.2% patients. Mean dose of lacosamide was 296.88 mg/day. (Median dose 300mg/day)

## SEIZURE CONTROL PROFILE

87.5% of the study subjects had no seizures at 6 months of follow up. In the remaining 12.5% of patients, reduction in seizure frequency was observed. The difference in seizure frequency before and after lacosamide administration is statistically significant (P<0.001). Engel’s seizure score showed rapid decline following introduction of lacosamide. After 3^rd^ month it became low and remained steady, showing excellent seizure control.(Figure 1) The difference between initial and final Engel’s score is statistically significant (p< 0.001).

**Figure 1.**
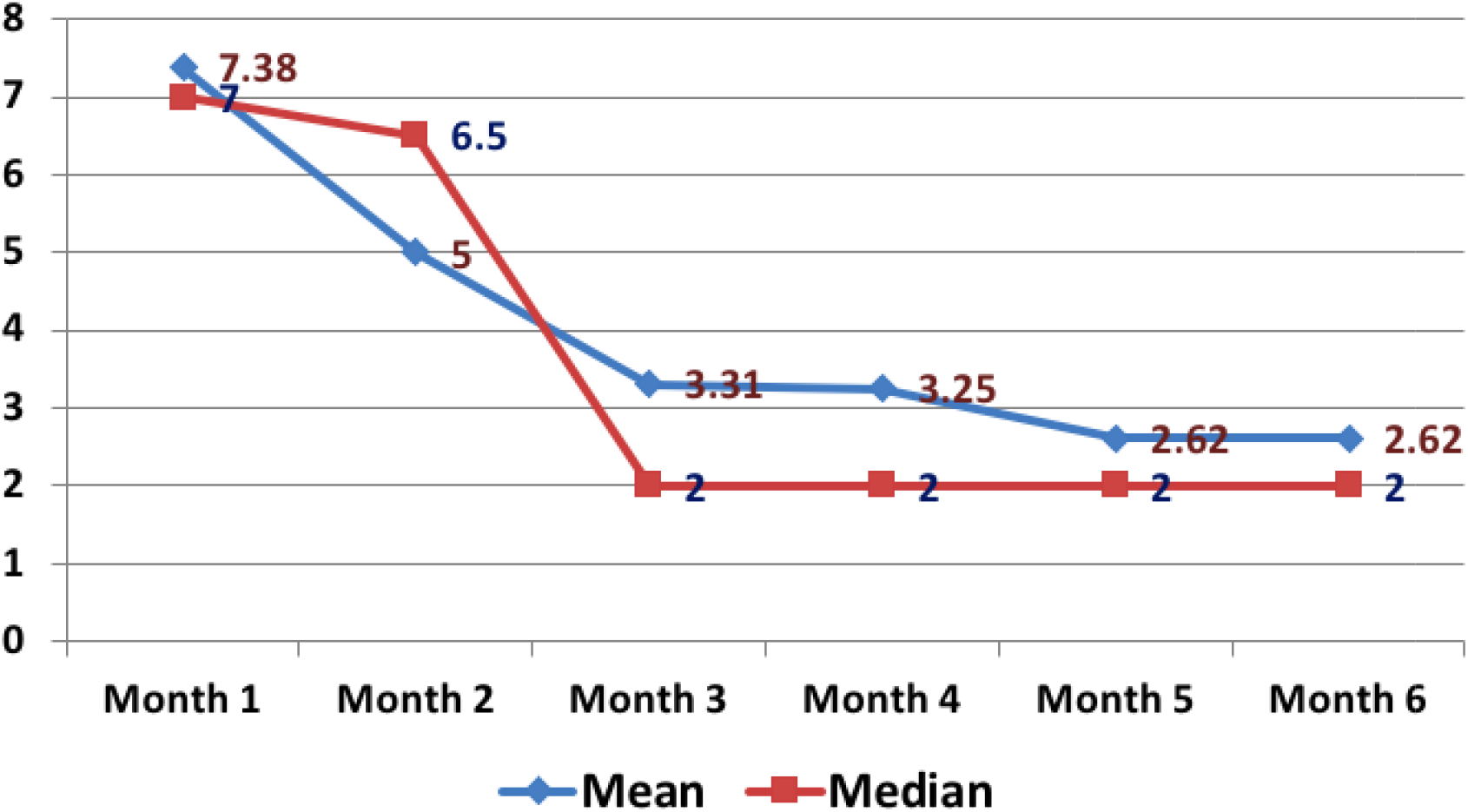
Engel’s Seizure score during the follow up period.

## BASELINE COGNITION

In the baseline evaluation, majority of the patients showed cognitive impairment. Difference in mean M-ACE total score from population normal [88] was statistically significant. No significant association was observed between baseline M-ACE scores with most of the socio demographic and clinical variables studied.

Though not statistically significant, it was observed that as the age advances cognition decreases. Orientation showed a **positive correlation** with years of schooling (p<0.05).

Of the 22 subjects, only 72.7% were able to perform Trail making test-part A. **Mean baseline time taken to complete the test was 78.4 seconds**. Difference in mean time taken by subjects from population average (29 seconds) was statistically significant (p<0.05). Mean score of digit span backward was 2.1, which was **less than normal** (4±1 depending on age and intellectual abilities).

No significant association was observed between baseline TMT-A, digit backward, digit symbol and block design test scores with most of the socio demographic and clinical variables except **TMT-A with education status (P 0.018) and Block design with Occupation. (P 0.021)**

In this study, it was observed that, base line trail making test Part-A (TMT-A), digit backward and digit symbol tests are correlated with schooling (p<0.05).

## PRE AND POST LACOSAMIDE COGNITION

Difference in pre and post lacosamide M-ACE total scores was not significant (p 0.139).(Figure 2) Difference in **Memory** showed statistically significant improvement (P 0.007). **Naming showed statistically significant improvement** (P 0.037).

**Figure 2.**
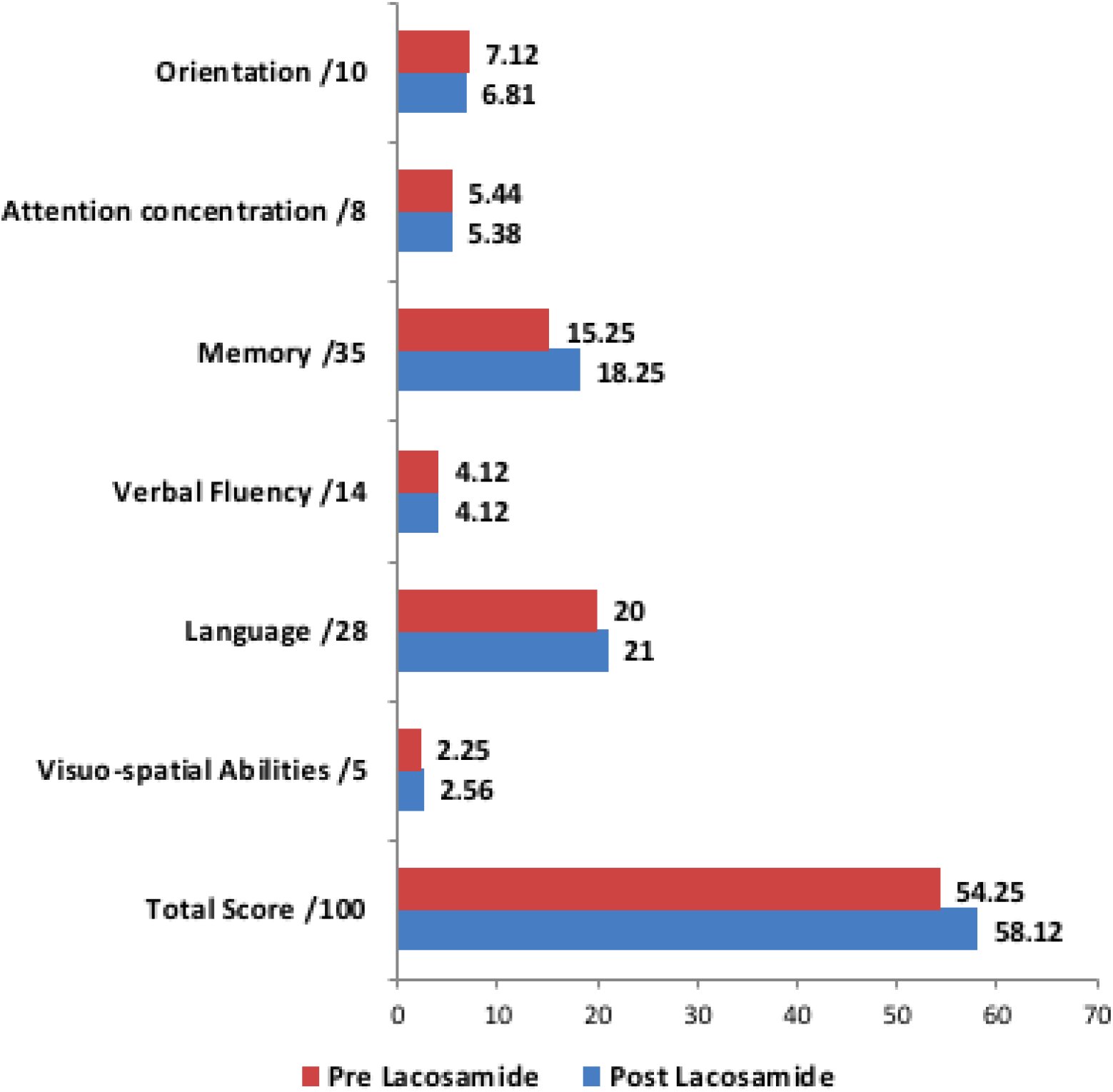
Clustered bar diagram comparing pre lacosamide and post lacosamide M-ACE scores.

In this study, only 31.25% subjects showed a reduction in M-ACE total score from the baseline value.(Table1) Maximum number of people showed deterioration in sub-domain orientation 37.5% and minimum in memory and visuo-spatial abilities 18.75% each.(Figure3)The association between number of AEDs and attention/concentration was statistically **significant** (p 0.03). Subjects from higher **socio-economic status** were having better improvement in total cognition scores **(P 0.03)** and sub-domain language (**P 0.03**) compared to subjects from low socio-economic background.

**Table 1:**
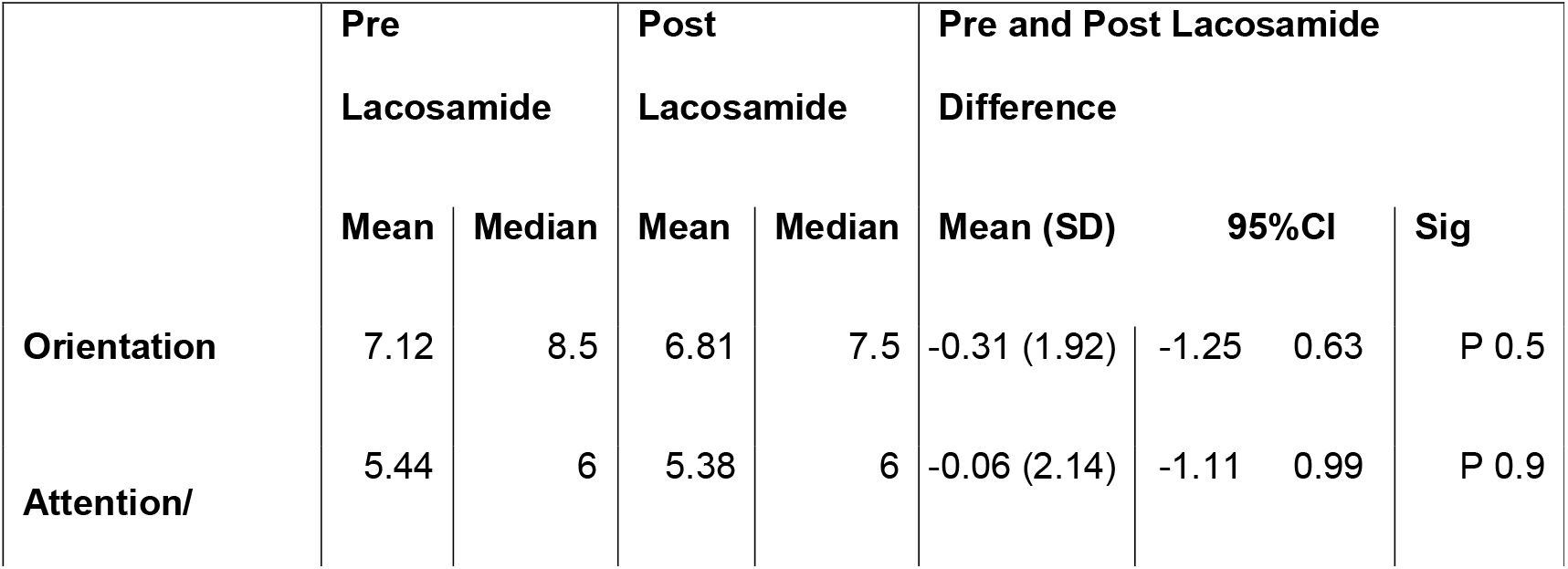
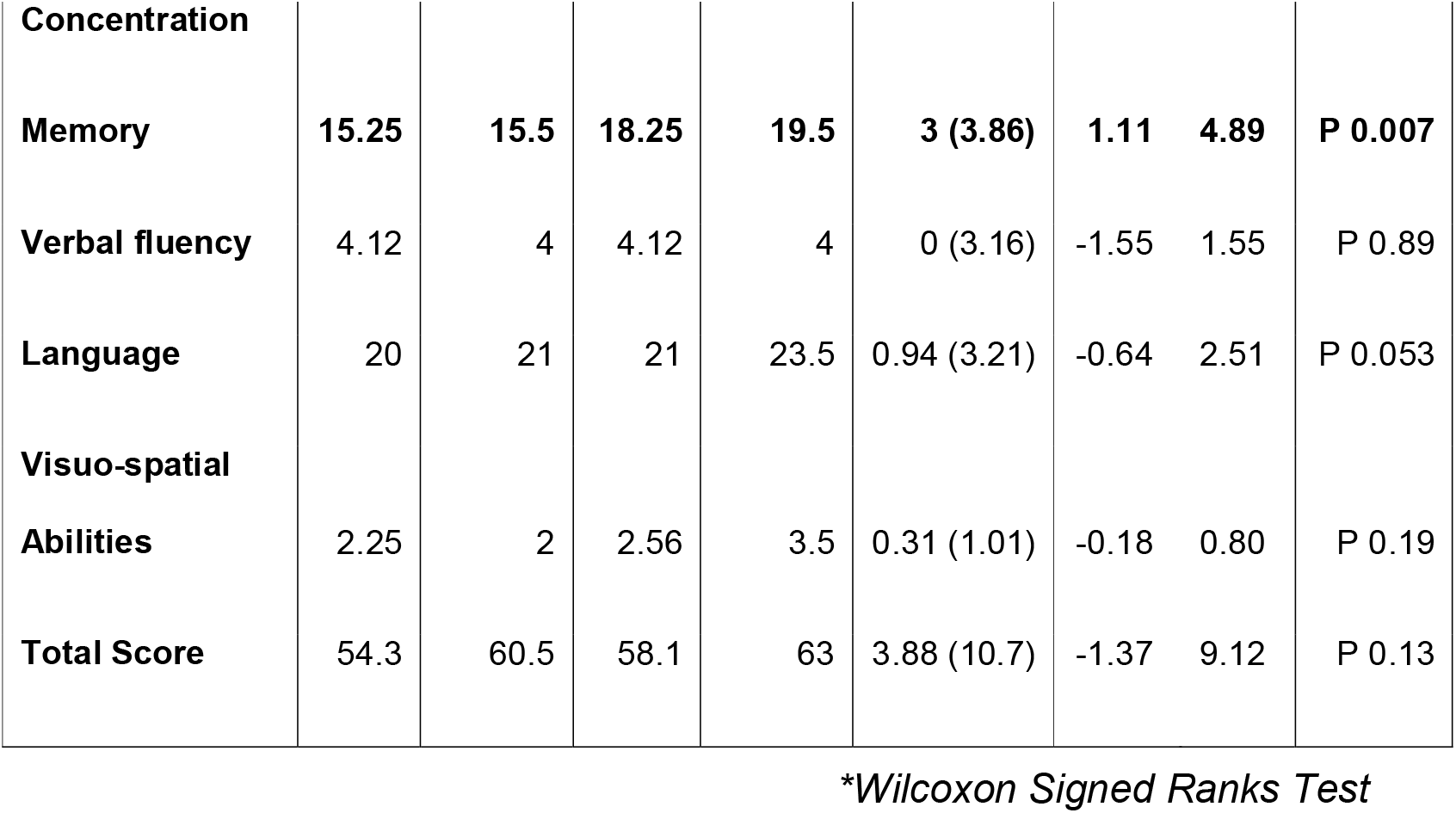
Pre lacosamide and post lacosamide M-ACE scores and their significance.

Though not significant, it was observed that as the age advances cognition decreases. Increase in number of AEDs and dose of lacosamide decreases cognition scores.

Negative correlation was shown by attention /concentration, memory, verbal fluency and total score with number of AEDs. **Difference in pre and post lacosamide language score showed statistically significant positive correlation with years of schooling**. Subjects getting four AEDs showed deterioration in post lacosamide M-ACE total scores compared to those who got three or two AEDS.

**Though not statistically significant**, it was observed that mean time taken to perform TMT-A (frontal/executive functions) was higher in post lacosamide subjects, which is an adverse cognitive response.

Post lacosamide **digit backward** test scores (which assess **attention and working memory**) showed **significant** improvement from baseline scores (P 0.04). Digit symbol (which measures psychomotor speed and sustained attention) and block design tests (which measures spatial perception, visual abstract processing, and problem solving) showed improvement, but not significant. In this study, 25% of subjects showed post lacosamide deterioration in TMT-A performance and only one subject showed deterioration in digit backward test.

**Difference in TMT-A showed association with marital status** (P 0.042) **and educational status of the subjects** (P 0.019). **Digit backward test also showed association with occupation status of the subjects** (P 0.047). It shows that subjects with higher education took lesser time for completing TMT-A in post lacosamide situation.

Significant association was observed between **CT/MRI abnormalities Vs. TMT-A** (P 0.016). **EEG abnormalities** showed significant association with **Digit Symbol** (P 0.03) **and Block design** (P 0.042) tests.

All the pre and post lacosamide cognition scores showed statistically significant positive correlation in this study. (Table 2, Table 3)

**Table 2:** Correlation between differences in pre and post lacosamide cognition scores (M-ACE)

| **Pre Vs. Post lacosamide scores** | **Spearman's rho** | **P value** |
| --- | --- | --- |
| Orientation | 0.774 | <0.001 |
| Attention Concentration | 0.506 | 0.045 |
| Memory | 0.603 | 0.013 |
| Verbal Fluency | 0.19 | 0.47 |
| Language | 0.85 | <0.001 |
| Visuospatial abilities | 0.88 | <0.001 |
| Total score | 0.78 | <0.001 |

**Table 3:** Correlation of pre and post lacosamide cognition scores (Trail making test-part A, digit backward test, digit symbol test, block design test)

| **Pre Vs. Post lacosamide scores** | **Spearman's rho** | **P value** |
| --- | --- | --- |
| **Trail making test part-A** | 0.666 | 0.025 |
| **Digit backward** | 0.822 | <0.001 |
| **Digit symbol** | 0.889 | <0.001 |
| **Block design** | 0.496 | 0.051 |

It was observed in this study that **as age advances, time taken to complete TMT-A increases, which is an adverse cognitive effect**. Though not significant, number of AEDs and dose of lacosamide were negatively associated with pre and post lacosamide difference in most of the cognition scores.

## DISCUSSION

Several interacting factors including age of onset, seizure (type, duration, frequency, severity & etiology), hereditary factors, psychosocial issues and cognitive side effects of antiepileptic drug (AED) therapy contribute to cognitive dysfunctions in epilepsy.[14] AEDs control seizures by causing global changes in the excitation levels in central nervous system. These alterations may result in cognitive deficits. Cognitive dysfunctions can negatively affect tolerability, compliance, and long-term retention of the treatment and can significantly affect every day functioning and quality of life of the patient. This study was aimed at assessing the effect of lacosamide on cognitive functions in patients with localization related epilepsy attending the neurology department of a tertiary care hospital.

In this study, lacosamide was discontinued by 27.2% patients; of whom 22.7% discontinued the drug themselves giving reasons such as dizziness (13.5%) and financial burden (9%). Nunes et al observed that discontinuation rate has been seen to be higher in lacosamide treated patients when compared to other AEDs.[15] Flores et al observed that 38% patients discontinued lacosamide, of which 20.8% withdrew due to intolerable ADRs alone. [16]

87.5% of the study subjects had no seizures at 6months of follow up. In the remaining12.5% of patients, reduction in seizure frequency was observed. In patients who had secondary GTCS, the seizure type changed to complex partial seizures. If a patient who used to experience GTCS earlier is experiencing only SPSs and CPSs later on, then it might be a sign of improved control of the seizures. In the present study, all patients reported improvement in seizure severity.

The difference in seizure frequency prior to lacosamide administration and post lacosamide is statistically significant (P <0.001). This goes in accordance with various similar studies, where the median percentage reduction in seizure frequency per 28 days from baseline to the maintenance period for lacosamide ranged from 32.7% to 35.3% for 200 mg/day and 36.4% to 41.1% for 400 mg/day. [2,8,13]

Engel’s seizure score showed rapid decline following introduction of lacosamide. After 3^rd^ month it became low and remained steady, showing excellent seizure control. **The difference between initial and final Engel’s score is statistically significant (p < 0.001)**.

Among the 22 patients observed in this study, 10 patients (45%) complained of dizziness. In a similar study by Flores et al, 48.7% patients reported adverse effects. Sedation and dizziness were the most common ADRs followed by nausea.[16] The incidence of dizziness due to lacosamide in various studies range from 10.4% to 44.5%.[17,18,13,2] It was observed that median dose of Lacosamide was equal (300mg/day) among both groups. **Adverse events were not seen to be dose dependent in this study**. But in many studies, adverse events were observed to be dose dependent. [19]

Though **not** significant, those who were concomitantly receiving sodium channel blocking AEDs in this study had higher frequency of dizziness compared to subjects receiving non sodium channel blocking AEDs. This is in accordance with the study by Novy et al, in which lacosamide when concomitantly administered with other sodium channel blocking AEDs was observed to cause increased side effects (diplopia, dizziness, drowsiness).[20]

In the present study, comparison of on-treatment versus baseline ECG results did **not** demonstrate any change in heart rate, PR interval, QT interval, or QRS duration in the study subjects. **No** cardiac adverse events were reported during the study period. Cardiac adverse effects including dose dependent PR interval prolongation, first and second degree AV block and atrial fibrillation/flutter has been observed with lacosamide in few studies. [21,22,23]None of the subjects in the current study had any cardiac conduction problems or were taking drugs known to cause PR interval prolongation or had severe cardiac disease such as myocardial ischemia or heart failure at base line. This factor might be the reason for the lack of cardiac adverse events observed in the present study.

A total of 3 patients (13.5%) withdrew from this study due to dizziness experienced during treatment with lacosamide. In various studies, percentages of withdrawals during the lacosamide treatment period because of adverse effects are 8%, 8.7%, 17% [2,13,24] etc. with dizziness, ataxia, nausea, vomiting, diplopia etc. being the most common reasons.[25] Causality of ADR assessed by Naranjo’s Algorithm showed 100 % probable reactions.

It was observed in this study that the mean difference in pre and post lacosamide scores deteriorated in orientation and attention/concentration. Verbal fluency remained same in pre and post lacosamide assessment. Language, visuospatial abilities and total score showed improvement. Difference in **Memory** showed statistically significant improvement (P 0.007). **Recall and Retrograde memory showed statistically significant improvement**. (Recall P 0.02). Retrograde Memory (P 0.021)). **Naming showed statistically significant improvement** (P 0.037).

In a similar study, Helmstaedter observed that 9% of patients treated with lacosamide showed significant improvement in memory functions. (11)A meta analysis of 10 lacosamide randomized controlled trials for various indications observed that lacosamide is not associated with any significant cognitive adverse events. Memory impairment even though reported was found to be statistically not significant.[26]

The association between number of AEDs and attention/concentration was statistically **significant** (p 0.03).The observation in the present study is similar to the observation by Meador KJ that polytherapy (co-administration of multiple anticonvulsants) contributes to the risk of cognitive dysfunction and also increases the intensity of cognitive dysfunction. [27]

In this study, significant correlation was observed between pre and post lacosamide M-ACE total scores and all sub domain scores except Verbal fluency. (Table 2) Negative correlation was shown by attention /concentration, memory, verbal fluency and total score with number of AEDs. But years of schooling showed a positive correlation with M-ACE total score and most of its sub-domains. This goes in accordance with Meador KJ who observed that polytherapy (co-administration of multiple anticonvulsants) contributes to the risk of cognitive dysfunction and also increases the intensity of cognitive dysfunction. [27] This also goes in accordance with Mathuranath et al who observed that level of education is the demographic factor that significantly affects the M-ACE total score in Malayalam speaking population in southern INDIA. [28,29]

Post lacosamide **digit backward** test scores (which assess **attention and working memory**) showed **significant** improvement from baseline scores (P 0.04). In this study, 25% of subjects showed post lacosamide deterioration in TMT-A performance and only one subject showed deterioration in digit backward test. This goes in accordance with Helmstaedter et al. who observed that 23% of patients treated with lacosamide showed significant improvement in executive functions and 14% patients deteriorated in executive functions.[11]

All the pre and post lacosamide cognition scores showed statistically significant positive correlation in this study. (Table 3) Of the subjects with EEG abnormalities, 50% showed deterioration in psychomotor speed and sustained attention from baseline while none with normal EEG deteriorated. Statistically **significant** (p 0.05).

**Difference in TMT-A showed association with marital status** (P 0.042) **and educational status of the subjects** (P 0.019). **Digit backward test also showed association with occupation status of the subjects** (P 0.047). It shows that subjects with higher education took lesser time for completing TMT-A in post lacosamide situation.

Significant association was observed between **CT/MRI abnormalities Vs. TMT-A** (P 0.016). **EEG abnormalities** showed significant association with **Digit Symbol** (P 0.03) **and Block design** (P 0.042) tests. This goes in accordance with Hermann et.al. who observed that baseline volumetric abnormalities are predictive of an increased risk of a progressively abnormal cognitive course. [30]

In this study, while looking at correlation, all tests except digit symbol showed a positive correlation. It was observed in this study that **as age advances, time taken to complete TMT-A increases, which is an adverse cognitive effect**. This goes in accordance with Tombaugh et al and Ashendorf et al who have observed that TMT-A performance declines with increasing age but not with education. [31,32]

Multiple factors might have played a role in the improvement in cognitive functions observed in this study. One factor can be the significant reduction in seizure frequency. It is a proved fact that continuing seizures contribute to cognitive deterioration in epileptic patients by inducing hippocampal sclerosis. [33] The reduction in seizure severity might also be an important contributor to the cognitive improvement. Previous studies have shown that severe seizures and status epilepticus reduces cognition. [34–36]

Another important factor might be the reduction in dose of or stopping of other concomitant AEDs which are associated with cognitive deterioration (phenobarbitone, carbamazepine etc.)

### Limitations

This study was conducted as a prospective observational study. For better assessment of association an analytical study, preferably a randomized, controlled trial is desirable. Another important limitation was the small number of subjects included in this study. This was because lacosamide is a comparatively new drug and so it is being prescribed only to patients in whom treatment with first line AEDs fail. Another limitation was the restricted period of monitoring of adverse drug reactions. For complete assessment of adverse effects of lacosamide on cognition, patients have to be followed up over a longer time.

### Conclusion

Excellent seizure control is observed in patients with refractory localization related epilepsy treated with lacosamide. Also, lacosamide has no serious adverse effects or drug interactions. In the above study, it is observed that unlike many AEDs, lacosamide contributed to significant improvement in cognition and can improve the quality of life in such patients.

## Data Availability

No external datasets or supplementary material online

## Declarations

### Availability of data and material

‘Not applicable’

### Code availability

‘Not applicable’

### Authors’ contributions

Study conception, design and material preparation: Dr Sophia B Modi, Dr Thomas Iype and Dr Asha S

Data collection : Dr Sophia B Modi, Dr Thomas Iype and Mrs Reeja Rajan.

Formal data analysis: Dr Libu GK

Writing – original draft: Dr Sophia B Modi

Writing – review & editing: Dr Asha S, Dr Sophia B Modi, Dr Thomas Iype

All authors read and approved the final manuscript.

### Consent to participate

Informed consent was obtained from each subject / guardian/ relative

### Consent for publication

Informed consent was obtained from each subject / guardian/ relative

